# Management Algorithm of External Fixation in Lower Leg Arterial Injury for Limb Salvages

**DOI:** 10.1101/2021.03.01.21252666

**Authors:** Lei Jin, Song Zhang, Motao Liu, Yuxuan Zhang, Xin Lin, Dehong Feng, Kejia Hu

**Author notes:** D. Feng and K. Hu are the Co-Correspond authors of this work. L. Jin and S. Zhang and contributed equally to this work.

## Abstract

**Purpose:** The purpose of this study was to review the roles of using external fixation to rescue the patients who sustained arterial injuries in the lower legs.

**Methods:** Demographics, surgical treatment and outcomes in 88 patients with lower leg arterial injuries treated by external fixation at two trauma centers from 2009 to 2018 were reviewed. The primary outcome was the rate of successful lower leg salvage, while secondary outcomes were complications and functional recovery.

**Results:** 80 patients (90 legs) maintained a successful lower leg salvage. The patients were followed up for an average of 15.5±5.5 months. 6 patients (8 pins) experienced pin-tract infection, pins loosening happened in 2 patients (4 pins), 7 patients (7 legs) developed wound superficial infection, 3 patients (3 legs) with a deep infection developed osteomyelitis, 16 patients (17 legs) suffered the bone nonunion or bone defect. The average healing time of fracture was 5.6±4.3months. The maintain of external fixation average time was 5.8±3.6 months.

**Conclusion:** With correctly judging the condition of limb ischemia, mastering reasonably the operation indications, and preventing complications, good clinical effects can be achieved when external fixation is used.

**Level of evidence:** Retrospective cohort, level IV.

## Introduction

Limb loss following lower leg arterial injury is common and has serious implications for the patient’s life and functionality. The lower leg arterial injury is sometimes accompanied by comminuted fracture, severe wound contamination and damage to skin, soft tissue and nerve. It can easily lead to compartment syndrome, bone exposure infection, nonunion, and invasive infection, which can be reasons for limb loss, even life-threatening. Previous study indicated that ischemia time of more than 6 h was associated with increased rates of limb loss, also there is no question that amputation rates climb as ischemia times lengthen. So prompt diagnosis and management to avoid irreversible soft-tissue ischemia with consequent reperfusion injury, which may necessitate amputation^1,2^. A suitable skeletal stabilization can be extremely helpful in the patient with a severe or dysvascular extremity injury^3^.

Technological characteristics of external fixation can improve the condition of the limb to sustain it from the acute phase to the reconstructive phase with few disadvantages. This benefit is especially evident in simplifying operation methods and reducing operating time. The use of external fixation can yield excellent stability to allow the vascular repair to be performed in a controlled environment to protect the completed vascular repair from disruption^4^.Furthermore, external fixation can be adapted to fitting the uniqueness of the individual patient, and can minimally disturb the local soft tissues, which creates a good environment for initial recovery of function.

The literature which specifically focused on the treatment with external fixation and outcomes of the lower-limb vascular injury was limited. The purpose of this study is to investigate the limb salvage outcome and functional results of these limb-threatening injuries through external fixation treatment.

## Materials and methods

From January 2010 to December 2016, trauma patients with lower leg arterial injury which surgically treated with external fixation by the Microsurgery team in two level 1 trauma centers (Wuxi People’s Hospital and Wuxi Orthopedic Hospital) were retrospectively included. This study was performed with institutional review board approval in accordance with the Helsinki Doctrine.

Two separate reviewers performed the data collection, recording patient demographics, medical comorbidities, injury mechanism, Gustilo–Anderson classification^5^, mangled extremity severity score (MESS)^6^, injury severity score (ISS)^7^, AIS abbreviated injury scale^8^, time to surgery, flap use in soft-tissue reconstruction, follow-up time, and postoperative complication. The primary outcome of interest was lower leg salvage. Secondary outcomes included complications and functional recovery. And we use the Lower Extremity Functional Scale (LEFS) to evaluate the functional recovery of lower limbs, the Visual Analogue Scale (VAS) and the Quality of Life Scale (QOL) to evaluate pain and life quality correspondingly.

Statistical analyses were performed using Stata version 14.0 MP (StataCorp) to assess for differences in patient demographics, injury characteristics, treatment course, and complications. The Kolmogorov-Smirnoff test was used to test whether the data were normally distributed. Normally distributed data were expressed as a mean ± standard deviation, and skewed data were expressed as median (interquartile range). F-test was used for homogeneity of variance, independent samples t-test for an equal variance, and the non-parametric test was used for unequal variance. P<0.05 was considered statistically significant.

## Results

### Patients Demographics

88 patients (98 legs) were diagnosed with a lower leg arterial injury with an unstable bone fracture or knee dislocation, which surgically treated with external fixation was included in the study. Table 1 presents the demographic data for the study population: age, gender, and injury mechanism.

**Table 1.** Demographics of patients with lower leg arterial injury.

| Demographics | Case (N) |
| --- | --- |
| **Age, years, mean ± SD** | 32.7±10.8, (range, 16-65) |
| **Gender** |  |
| Males | 72 |
| females | 16 |
| **The mechanisms of injury** |  |
| Motor vehicle accident trauma | 40 |
| Bruise injury caused by heavy objects | 34 |
| Falling injury | 3 |
| Twist injury by working machines | 8 |
| Cutting injury | 1 |
| Explosion injury | 2 |
| **The types of arterial injury** |  |
| Popliteal artery | 35 (36 legs) |
| Anterior tibial artery | 16 (20 legs) |
| Posterior tibial artery | 17 (20 legs) |
| Anterior and posterior tibial artery | 20 (22 legs) |
| **The types of skeletal fracture** |  |
| Upper 1/3 tib/fib | 40 (44 legs) |
| Middle 1/3 tib/fib | 18 (22 legs) |
| Distal 1/3 tib/fib | 28 (30 legs) |
| Traumatic knee dislocation | 2 (2 legs) |
| **Soft tissue injury** |  |
| Open (Gustilo IIIc) | 80 (88 legs) |
| Closed | 8 (10 legs) |
| **Injury evaluation** |  |
| AIS score, mean ± SD | 10.1±1.0, (range, 9-12) |
| MESS score, mean ± SD | 5.8±1.4, (range, 2-10) |
| ISS score, mean ± SD | 18.5±2.5, (range, 15-22) |
| **Injury to Surgery time, hours, mean ± SD** | 5.5±3.2, (range, 3-12) |

N number of patients, SD standard deviation, AIS abbreviated injury scale, MESS mangled extremity severity score, ISS injury severity score; Data were reported as the number (injured legs) of patients except where noted.

**Table 2.** Complications of lower leg arterial injury patients treated with external fixation.

| Complication | N | Treatment |
| --- | --- | --- |
| Pin tract infection | 6 (8 legs) | Effective antibiotics |
| Pins loosening | 2 (2 legs) | Exchanged the pins |
| Post-operative wound superficial infection | 7 (7 legs) | Effective antibiotics |
| Deep infection developed osteomyelitis | 3 (3 legs) | Thorough debridement, sequestrectomy and bone cement placement with vancomycin |
| Bone nonunion or bone defect | 16 (17 legs) | Bone grafting or bone transporting |

Patients suffered from lower leg atrial injuries with an unstable bone fracture or dislocation were classified into open injury and closed injury. Then, all the patients were treated with external fixation. And our study used Primary end-to-end anastomosis and Autologous Vein Graft to repair artery. Lastly, we classified the wound closure into different situation (Figure 1).

**Fig 1.**
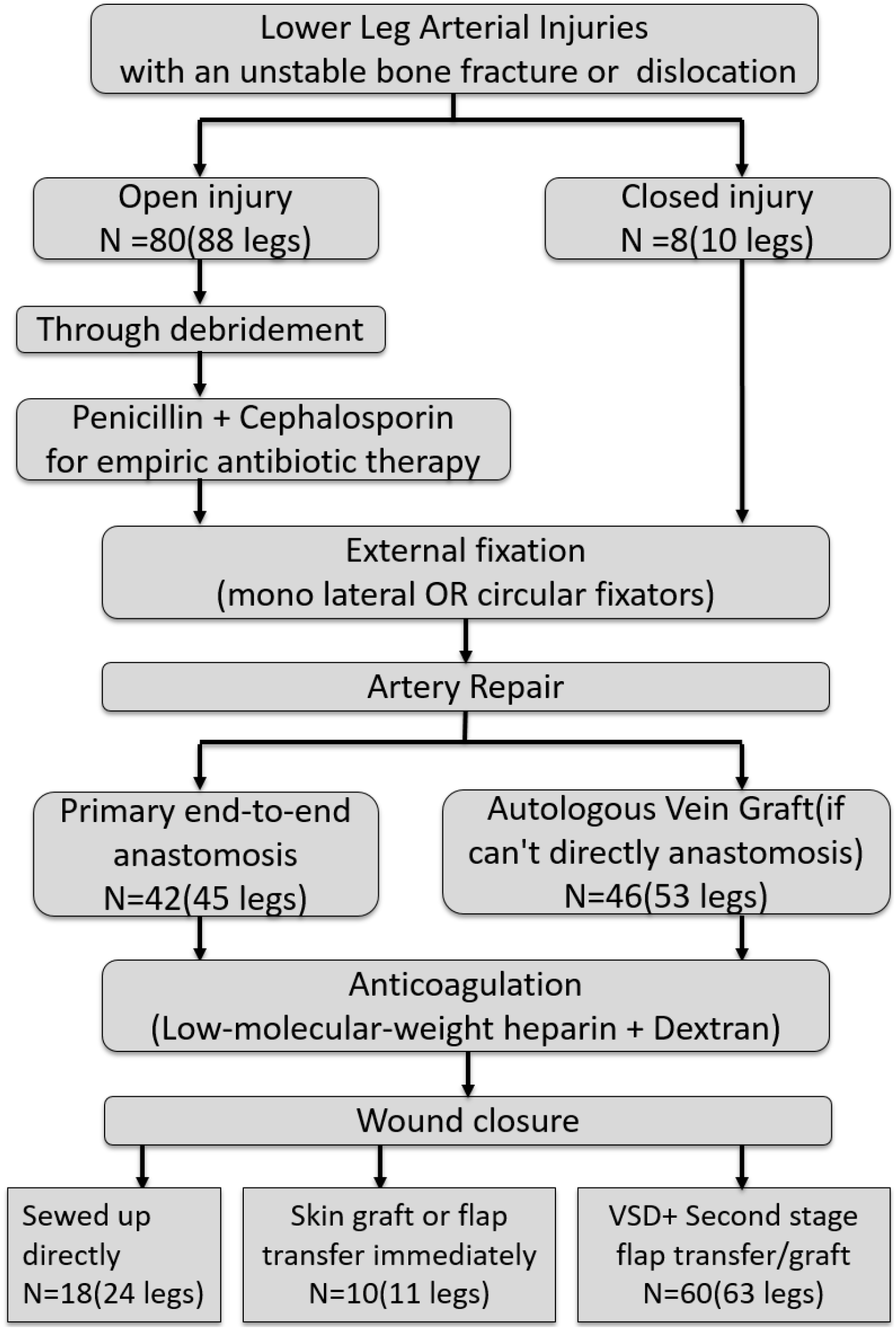
The flowchart of the application of external fixation in lower leg arterial injuries.

### Primary Outcomes

External fixation (including mono lateral and circular Fixators) was performed in all the 88 patients prior to arteries repair. We divided the patients into different parts by Gustilo classification. An example of the classification was presented in Figure 2.

**Fig. 2.**
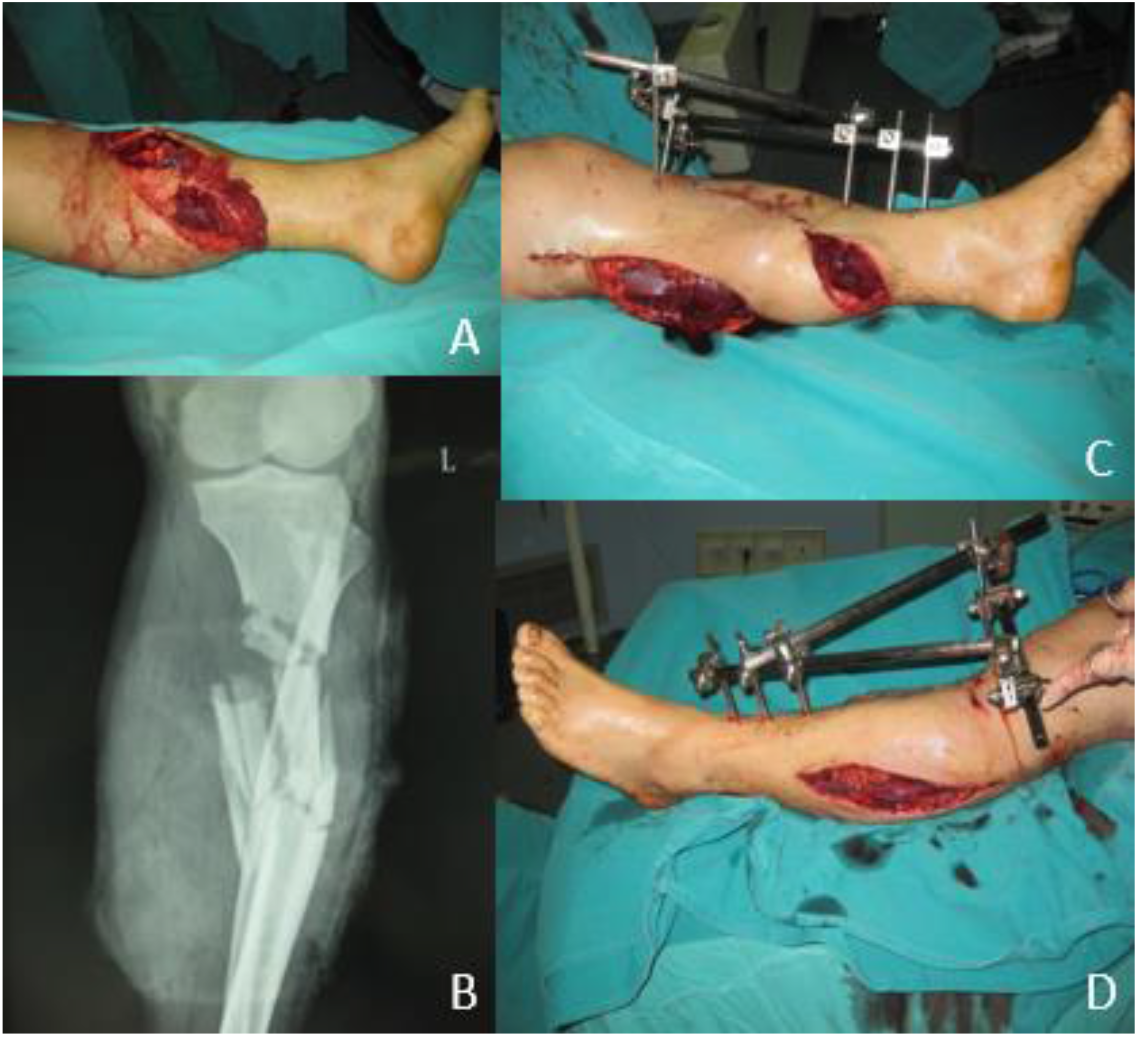
An example of the classification and the application of external fixation. (**A**) Patient was classified as type IIIc by Gustilo classification. (**B**) Preoperative radiograph showed an open comminuted fracture of the left tibia and fibula. (**C, D**) External fixation was used.

Primary end-to-end anastomosis under the microscope was preferred in 42 patients. Autologous vein graft from the contralateral leg was used in 46 patients due to the artery was shortened after thorough debridement (Fig 3).

**Fig. 3.**
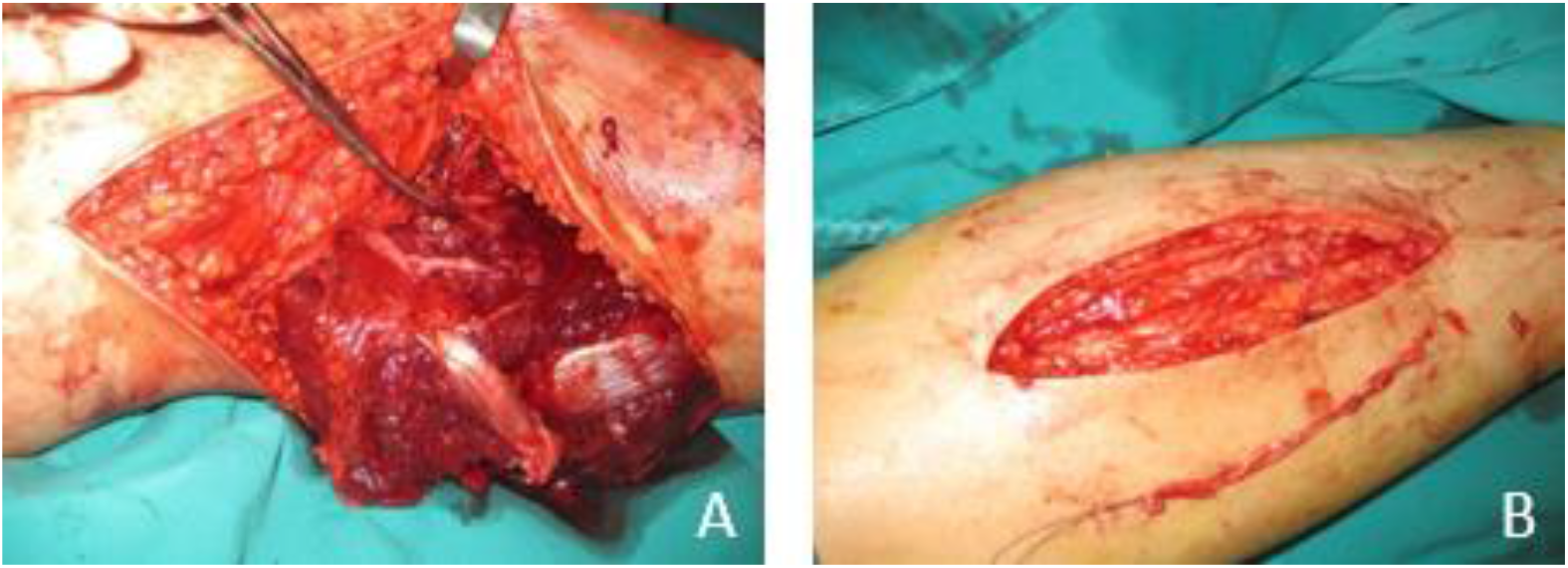
An example of the Autologous vein graft. (**A**) The popliteal artery was completely lacerated and shortened. (**B**) Autologous vein graft from the contralateral leg was performed.

**Fig. 4.**
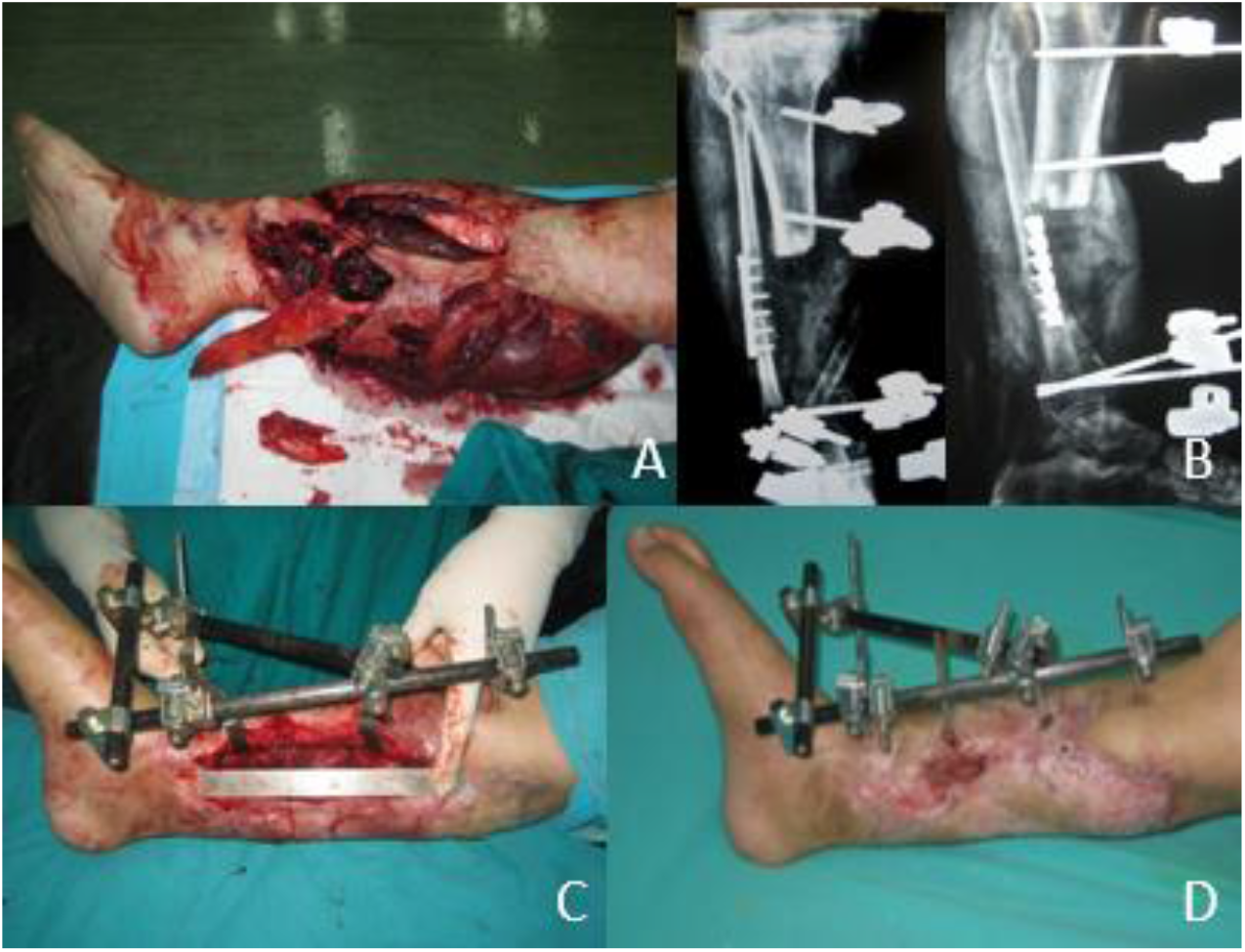
The use of external fixation and the outcome of the patient from Fig.3. (**A**) Patient was classified as type IIIc by Gustilo classification. (**B**) External fixation was used after vascular repair. (**C**) 12 months postoperative appearance after external fixation. (**D**) The tibia was shortened about 14cm.

**Fig. 5.**
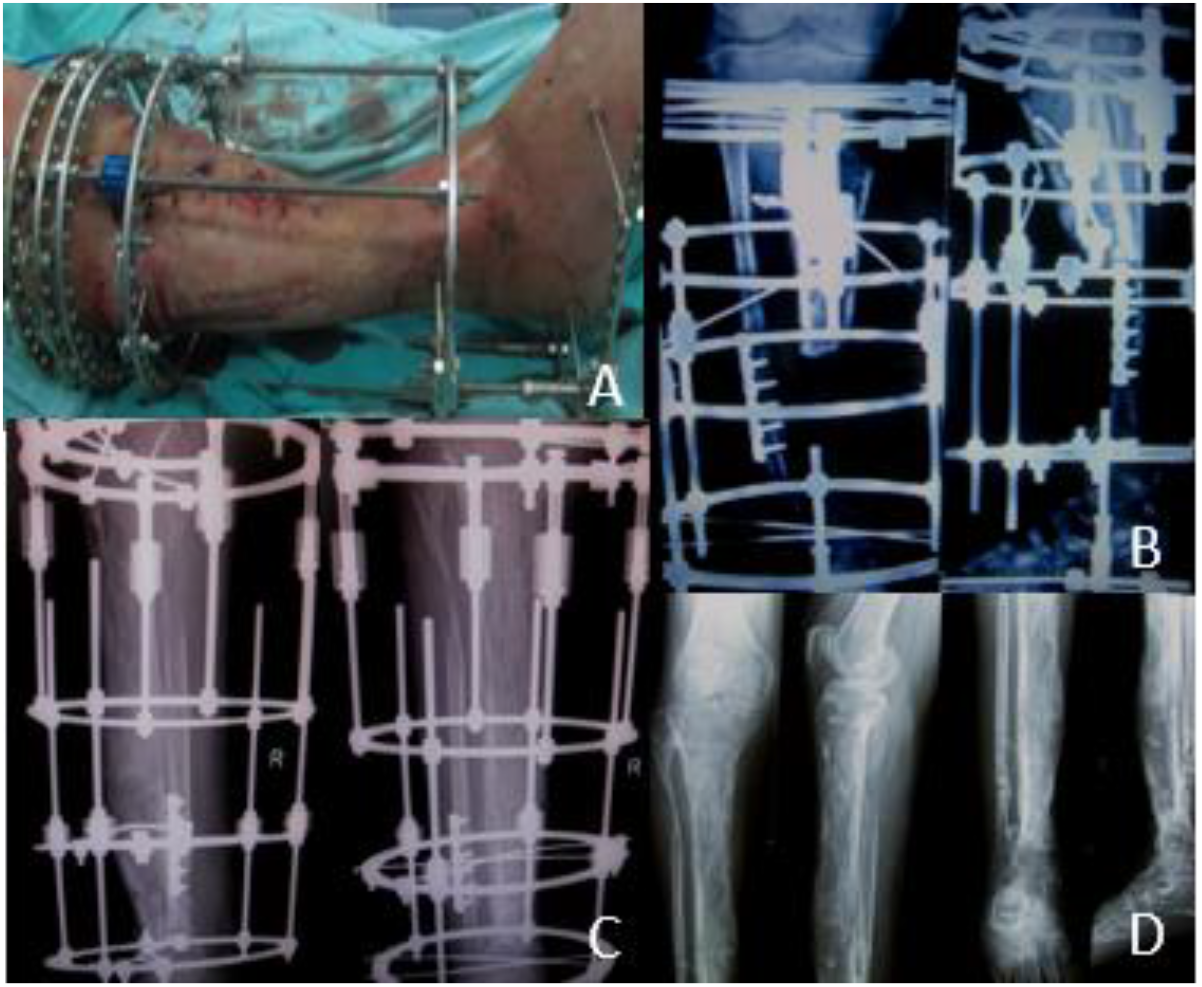
Imaging method was used to exluate the prognosis of the patients. (**A**) Most of wound was closed during shortening-lengthening operation. (**B**) Postoperative radiograph showed a good external fixation. (**C, D**) Anteroposterior and lateral X-ray film at 12 months after operation, and after the removal of fixators.

No patients died in this study, 8 patients (8 legs) underwent secondary amputation due to the onset of artery blood-circulation crisis postoperatively, no sign of revascularization after treated by spasmolytic medicine, or transplantation of the contralateral greater saphenous vein. All the 8 patients were in a poor general condition, who had a MESS score of 8 or above.

80 patients (90 legs) obtained successful limb salvage, and the salvage rate was 81.6%. 80 patients were followed up for 12 months to 3 years, with an average of 15.5±5.5 months. The external fixation time was 4 to 12 months, with an average of 5.8±3.6 months. The average healing time of fracture was 5.6±4.3 months, ranging from 3 to 13 months.

### Complications

Minor complications included pin-tract infection, pins loosening and wound superficial infection.

Three patients (3 legs) with a deep infection developed osteomyelitis, treatment involved thorough debridement, sequestrectomy and bone cement placement with Vancomycin, when the infection was quiescent as indicated by inflammatory parameters at least 1 year later, removal of External fixation and bone grafting with internal plating fixation was performed. All of them got bone union postoperatively in 7 to 15 months, with an average of 10.6±2.1 months.

16 patients (17 legs) suffered bone nonunion, 6 to 12 months after wound healing, patients were performed removal of External fixation, internal plating fixation replacing and bone grafting or bone transporting, all of them got bone union postoperatively in 5 to 14 months, with an average of 9.2±3.2 months. Among them, 5 patients (5 legs) had unilateral limb shortening experienced surgery. (Fig.4) The length of the tibial bone defect was 4.5-14.0 cm with an average of 7.2±3.8 cm.

Limb shortening-lengthening method using Ilizarov technique (external circular Fixators) was applied, at the time of the latest follow-up, all patients had shortened length less than 2 cm. (Fig.5)

### Functional recovery

The LEFS was used to evaluate the functional recovery of lower limbs with artery injuries in 80 patients (90 legs), while pain and quality of life were accessed by VAS and QOL scale separately. The survey measurements of the scales were shown in figure 6. All three scales tend to be improved over all three follow-up time points.

**Fig. 6.**
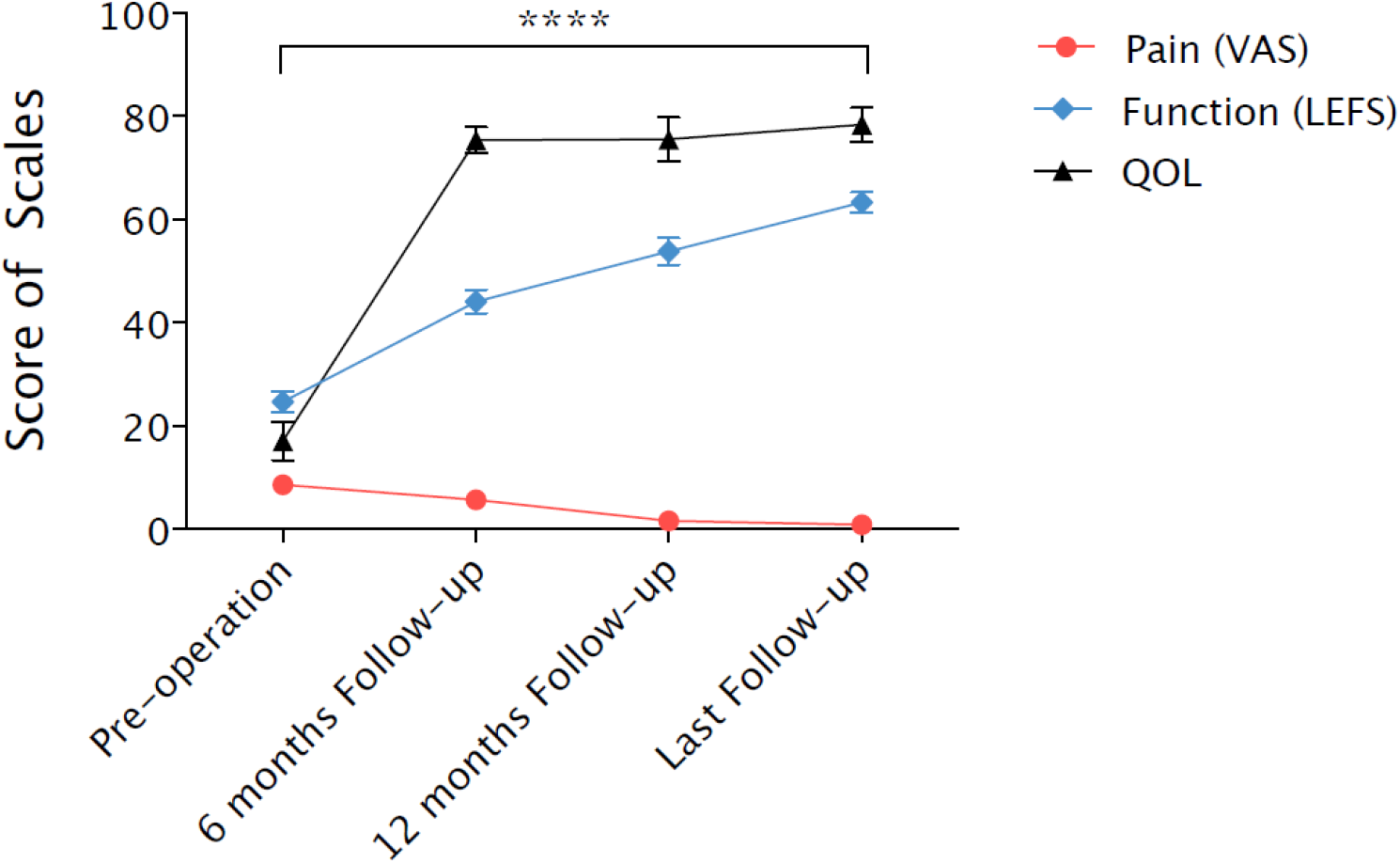
The scale of VAS, LEFS, QQL from the pre-operation to follow-up time. The VAS, LEFS and QOL score of the patient from Fig.4 was detected and statistical tests were conducted. We can see the tendency of the scores: the VAS score was decreased, whether the LEFS and QOL scores were increased over time. Statistical tests were conducted on the three groups of data, and it had statistical difference between group(pre-operation) and group(last follow-up).

**Fig. 7.**
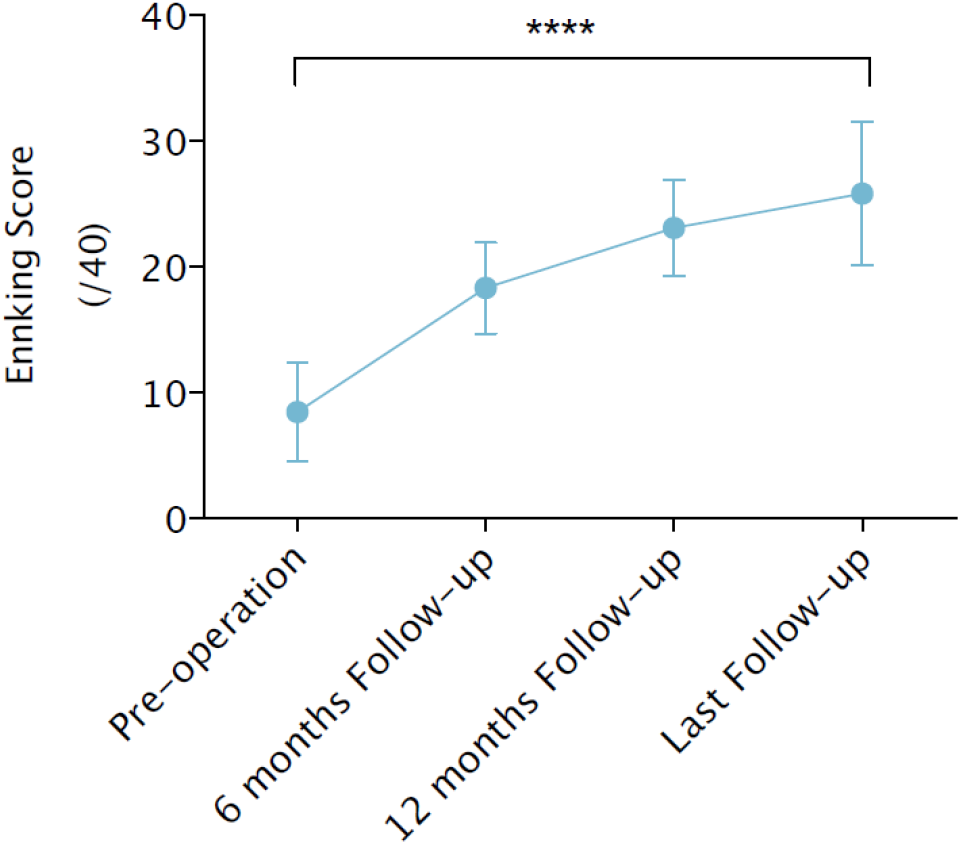
The Ennking score from pre-operation to follow-up time. The Ennking score was increased over time, and different follow-up time’s scores had statistical differences.

**Fig. 8.**
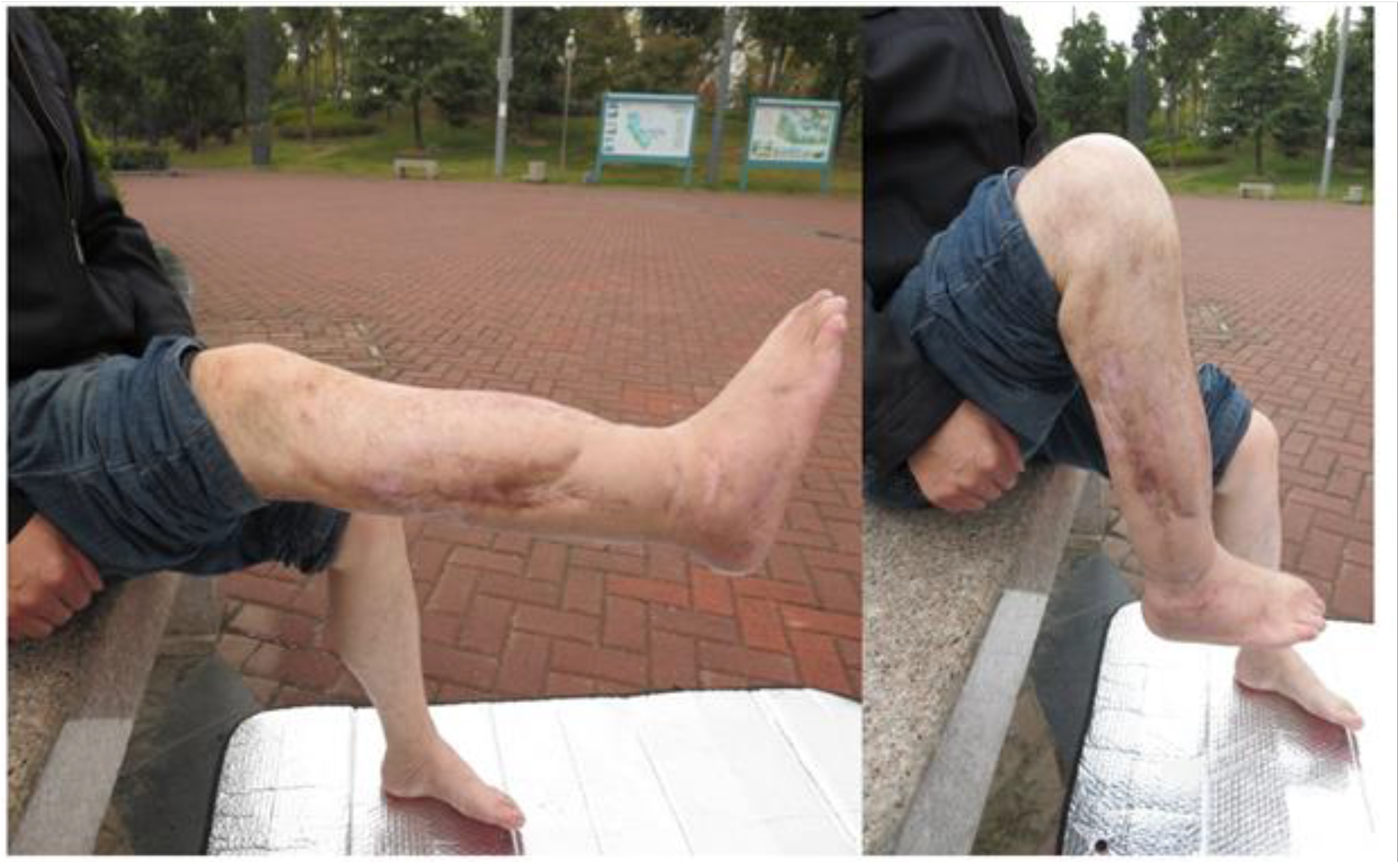
Patients showed in Fig.4 had equal limb length at 18 months postoperatively. The functions of the knee and ankle were good, with a Enneking score of 30.

The functional outcome of 70 patients (78 legs) following reconstruction with free tissue transfer or bony union was evaluated using the Enneking score system^9^. The latter is determined by clinical examination and based on an assessment of the degree of physical disability and psychological acceptance of the reconstruction. The Enneking score is expressed as a percentage of the non-injured contralateral limb and was measured routinely at the orthopedic clinics. Mean Enneking score for patients with vascular injury was 23.8±12.5, with a range of 7 to 38(Fig.7).

## Discussion

### Advantage and Indication

The use of external fixation is less invasive, can achieve adequate stability, and provide good access for wound management without compromising stability^10^. Vascular injuries of the lower legs, if not treated early and competently, are a major cause of limb amputation and even life-threatening, limb salvage is critically dependent on ischemic time^11^. In 1963, Malan and Tattoni^12^ introduced the concept of the 6-h rule for re-vascularization, and most authors used 6h as the definition of early intervention, however, skeletal muscle and nerve are, in fact, even more sensitive to ischaemia^13,14^. Glass et al^15^ in a Kaplan-Meier survival curve analysis, demonstrated that limb salvage begins to fall almost immediately the time when any further delay results in a rapid decline in survival, which begins at about 3-4 h. So, the time of re-vascularization should be as short as possible to minimize ischemia time and re-perfusion time, thus prevent potential necrotic changes and ischemia reperfusion injury, which is the key to limb salvage.

Reconstruction of the bone framework is one of the important steps in the surgery of vascular repair, and the surgical sequence has been the subject of debate. Some surgeons advocated arterial repair before fracture fixation suggests that this method minimizes ischemic time^16-18^. Others suggest that by performing skeletal fixation first, the requirements for vascular reconstruction can be better measured, and stabilization avoids iatrogenic disruption to the subsequent vascular repair^19^. Iannacone et al^4^ suggested that immediate external fixation allows the vascular repair to be performed in a controlled environment to protect the completed vascular repair from disruption. Our experience suggested that the use of external fixation technology to enable skeletal fixation prior to vascular repair has yielded satisfactory results in the treatment of vascular injuries of the lower legs. Also, researchers say that we should pay attention to the dynamization of fracture fixation to improve the fracture healing process. In the bone healing process, stabilizing elements of the fixator are removed at some time during the treatment leading to greater flexibility of the fixation and it gets good results.^20^

### The application scene of external fixation

Though external fixation has many advantages, we should also know that only when we use external fixation properly will we get satisfactory results. A prospective study of 59 patients with Grade II or III open tibial shaft fractures compared internal and external fixation shows that the rate and extent of complications are lower with external fixation though both methods yielded excellent results^21^. Another RCT shows that external fixation is a satisfactory method of treatment for fractures of the tibial plafond and is associated with fewer complications than internal fixation^22^. Through another RCT, Leung et al drew a conclusion that plate fixation combined with percutaneous pin fixation for the treatment of intra-articular fractures of the distal part of the radius is better than external fixation^23^. Other researchers such as P Pairon agrees with the idea that external fixation is a safe option for stabilisation of extremity lesions in the polytraumatised patient as well as in fractures with severe soft tissue damage. Even so, he also mentioned that stable polytraumatised patients who could not benefit from initial stabilisation with an external fixator should immediately be treated with a definitive osteosynthesis^24^. Our study suggests that we should choose the proper fixator in order to improve patients’ prognosis rather than use external fixator without taking patients’ situation into consideration.

Furthermore, another viewpoint we should consider. Zi-Chen Zhao demonstrates that combined fixation is an effective and safe alternative for management of open tibial diaphyseal fractures compared with external fixation^25^. In actual circumstances, we may choose the proper fixation method depended on types of fracture, ages of the patients et al.

### Complication

Pin tract infection is one of the most common complications of external fixation. Occurring in 10.3% of our patients is comparable to those of 9.4% to 30% reported by other studies^26,27^. Infection varies from minor inflammation remedied by local wound care; to superficial infection requiring antibiotics, local wound care, and occasional pin removal; to osteomyelitis requiring sequestrectomy. Higher rates of pin tract infection are seen when the pins are placed through large volumes of soft tissue (for example, thigh)^10^. So external fixator pins should be applied outside the zone of injury to span the zone of injury to minimize soft-tissue insult. Pins placed within the zone of injury are disadvantageous, which provided access for bacteria to invade potential space created by soft-tissue disruption, and their associated hematoma can cause deep infection. For different degrees of the pin tract infection, Checketts et al^28^ devised a classification system that aids in the formulation of treatment options.

Pin-bone interface loosening or failure mainly with bone resorption around needles, full weight-bearing too early, fracture gaps > 2 mm of unstable fracture, osteoporosis and so on. External fixation failure includes the pins and link rod crack and bending deformation. Repeating bending makes the metal fatigue, is a major cause of external fixation failure^29^.

Another common complication of external fixation is bone nonunion. Our study showed bone nonunion rate was 20% (16/80), considering that the original restoration is not satisfied, fracture gaps too large, severe soft tissue injury. Cross and Swiontkowski^30^ demonstrated as high as 13% bone nonunion rate using the external fixator, which was related to improper surgical technique, inaccurate fracture reduction, the initial severe trauma and the lack of elastic fixation changing in time. Menon^31^ demonstrated the early period static fixation, middle and later period elastic fixation could be beneficial to the bone union. The main reason for bone malunion is that the original restoration is not satisfied, the incidence of relative angular deformity was higher than rotational deformity, so the ideal fracture reduction should be performed as well as possible before placing pins, instead of excessive dependence on external fixator adjusting.

The general techniques for using external fixators are demanding, regardless of the specific fixator selected. Attention to detail is essential if the maximal advantage of the external fixator is to be gained and potentially serious complications are to be minimized. The initial treatment of the condition for which the external fixator is chosen must be considered first: irrigation, debridement, and reduction of the severe, open fracture; drainage, debridement, and sequestrectomy of the infected fracture. Furthermore, the surgeon must be familiar with the cross-sectional anatomy of the limb and with the relatively safe zones and danger zones for pin insertion. In addition, Hadeed demonstrated that two methods can be used to improve the stability of the pins. In the one hand, the operator should place pins closer to the fracture site, add more pins and increase the spread of the pins in order to improve the stiffness of the construct. In the other hand, he may increase the diameter of the rods or secure it closer in proximity to the bone^32^. By the way, the application of new materials of the pin such as hydroxyapatite-coated external will strongly enhance pin fixation.

### Management of arterial injuries

To recognize the vascular injury after surgical stabilization of trauma is vital for limb stability. Certain orthopedic injuries known to be associated with vascular injury are knee dislocation (especially posterior) and proximal tibial and fibular fractures, according to Treiman et al^33^, up to 23% of patients suffering knee dislocations had popliteal artery injuries. When making a diagnosis of vascular injuries, careful physical exam sensitive and specific, angiography should be used sparingly, the role of Color Flow Doppler is evolving. By the way, CT angiogram is recommended for use in the operation, for it has a high selectivity and specificity, and it is also noninvasive and requires less radiation^34^. Collateral arterial supply to the lower limb is frail and easily disrupted by soft tissue swelling or thrombosis, and thus the lower leg is almost totally dependent on the major trunk artery such as the popliteal artery, anterior tibial artery, and posterior tibial artery. An amputation rate of 16% following popliteal artery injury was reported by Hafez et al^35^, and Andrew et al^36^ demonstrated that the popliteal artery is vulnerable to injury from fractures or dislocations because it is tethered to both the distal femur by the adductor hiatus and the proximal tibia by the tendinous arch of the soleus muscle.

The key points of treatment are minimized preoperative delays, the primary end-to-end anastomosis is the first choice for Repair, if interposition grafts are required, auto logous vein graft is preferable to synthetic material, because a reversed saphenous vein graft from the contralateral limb has clearly superior patency rates, also can avoid foreign body in vivo^37-39^. Injury to veins should be repaired to minimize postoperative swelling, and compartment syndrome, which is a major risk factor for amputation following artery injury. There is evidence that fasciotomy done at the time of arterial repair, but before the development of compartment syndrome (prophylactic fasciotomy), may lower amputation rates^40^. Thrombosis is an important complication to suspect following arterial repair because it is both common and correctable. Heparinisation and routine monitoring of distal pulses postoperatively may reduce the incidence of thrombosis and allow for more rapid diagnosis when present.

## Conclusion

The external fixator has the advantages of reliable immobilization, can lower the possibility of the fracture or dislocation in a short time, which creates a good environment for vascular repair and shortens limb ischemia time. With correctly judging the condition of limb ischemia, mastering reasonably the operation indications, and preventing complications, the use of external fixator to treat lower leg arterial injury can obtain better clinical effects. Here, we provide a relatively standardized process by which clinicians can correctly classify the fracture in order to apply appropriate external fixation methods to improve patients’ prognosis.

## Data Availability

The data used to support the findings of this study are available from the corresponding author upon request.

## Acknowledge

We declare that we have no financial and personal relationships with other people or organizations that can inappropriately influence our work, there is no professional or other personal interest of any nature or kind in any product, service and/or company that could be construed as influencing the position presented in this article.

